# Equivalence of saliva RT-qPCR testing to nasal-throat/nasopharyngeal swab testing in the general practitioner’s setting to detect SARS-CoV-2

**DOI:** 10.1101/2021.09.30.21264181

**Authors:** Ilse Jonckheere, Liesbeth Faes, Yarah Overmeire, An De Vleeschauwer, Laura Vanden Daele, Nathalie Van Bruaene, Ilse Vandecandelaere, Britt Merlaen, Joannes van Cann, Jo Vandesompele

## Abstract

**Study design:** Saliva has been proposed as valid alternative for nasopharyngeal swab for RT-qPCR detection of SARS-CoV-2. The sensitivity is generally equivalent, and it comes with much less discomfort for the patient. While there is an overall good performance in the literature for adults, there is much less information on the use of saliva in children or in the general practitioner’s setting.

**Methods:** We tested a novel commercially available saliva collection kit with a virus inactivating and RNA stabilizing buffer (InActiv Blue^®^) in matched saliva and swab samples from 245 individuals, including 216 children, collected by general practitioners.

**Results:** Blind RT-qPCR testing of the saliva samples confirmed all 23 positives identified by swab testing (100% concordance), irrespective of age, presence of symptoms, or high-risk status. One child’s saliva sample was found low positive while negative on the nasopharyngeal swab, resulting in an overall relative sensitivity of RT-qPCR saliva testing of 104.3%.

**Conclusion:** Saliva collected in InActiv Blue^®^ can be a valid alternative for SARS-CoV-2 RT-qPCR testing in the general practitioner’s setting, including children.

## Intro

To control the COVID-19 pandemic, one needs to stop the spread of the SARS-CoV-2 virus by identifying and isolating infectious individuals. While a PCR test on a nasopharyngeal swab is generally considered to be the most sensitive diagnostic test, it comes with a few important shortcomings, such as discomfort for the patient (in particular, but not limited to children), the necessity for a trained healthcare professional to take a sample, risk for nosocomial virus transmission, and the identification of SARS-CoV-2 positive patients that are no longer infectious^1^. As COVID-19 is an airborne disease due to virus-laden aerosols expelled by an infectious individual^2^, several studies have evaluated saliva as an alternative and more easily accessible sample to detect SARS-CoV-2. In a meta-analysis, PCR testing on saliva yielded a sensitivity and specificity comparable to nasopharyngeal swab testing in ambulatory patients presenting with minimal or mild symptoms^3^. Given the ease of sample collection and increased patient comfort, the authors suggest that laboratories should consider adopting saliva as their first sample choice, especially in screening programs. In a more recent systematic review and meta-analysis, saliva PCR testing was specifically evaluated in children^4^. Comparable performance of saliva to nasopharyngeal samples was shown in both symptomatic and asymptomatic pediatric patients. While in general the RT-qPCR SARS-CoV-2 detection sensitivity and specificity on saliva is good, the various studies are quite heterogeneous in terms of patient inclusion criteria, volume of saliva collected, and saliva collection and preservation method. In our study, we aimed to evaluate a new saliva collection kit for self-sampling of a small volume of saliva under supervision using a virus inactivating and RNA stabilizing medium at the general practitioner’s office, with a focus on children. In total, matched swab and saliva was taken from 245 individuals, including 216 children.

## Study setup and results

In a first patient cohort, matched saliva and swab samples were collected by the general practitioner (GP) during visit of 209 children aged 5-16 years (median age of 9 years), because of high-risk contact and/or COVID-19 symptoms (May-June, 2021). While participants of the study were asked not to eat/drink/smoke/use chewing gum/candy/mint 30 min prior to saliva collection, and to rinse the mouth with water 10 minutes prior to saliva collection, this was not an inclusion criterium; it was noted which patients followed these recommendations.

Also, while children were asked to produce deep throat saliva (posterior oropharyngeal saliva) by scraping the throat, this was not an inclusion criterion if the participant could not produce such a sample. Upon collection, patient material was virus-inactivated and RNA-stabilized by InActiv Blue^®^ medium and picked up by medical lab 1 for routine RT-qPCR testing of SARS-CoV-2 on the nasopharyngeal or combined nasal/throat swab sample. Eleven swab samples (5.26%) were tested positive; these 11 positive and 62 (of the 198) randomly selected negative patients were sent for blind analysis to lab 3 using a validated RT-qPCR testing procedure for saliva (Figure 1).

**Figure 1:**
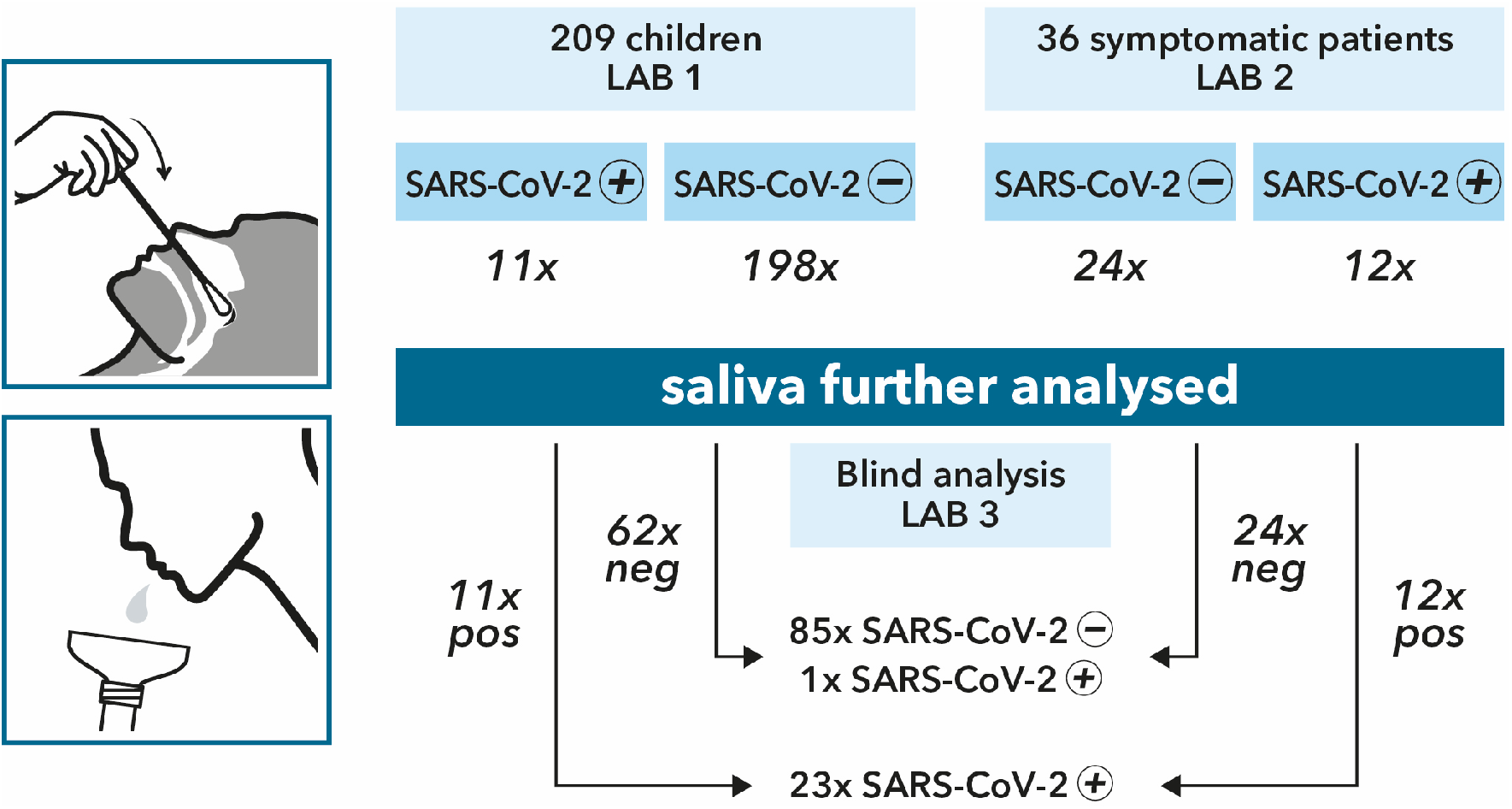
STARD diagram displaying study setup of matched saliva and nasopharyngeal or nasal/throat swab sampling of 245 patients at the general practitioner’s setting, including 216 children (209 from lab 1 and 7 from lab 2).

A second cohort of 36 symptomatic individuals (including 7 children) was selected by other GPs for matched sampling of saliva and nasopharyngeal swab, using the same instructions as described for the first cohort (May-June, 2021). Upon collection, patient material was virus-inactivated and RNA-stabilized by InActiv Blue^®^ medium and picked up by medical lab 2 for routine RT-qPCR testing of SARS-CoV-2 on the swab sample. Twelve swab samples (33%) were tested positive; all 36 saliva samples were sent for blind RT-qPCR analysis to lab 3 (Figure 1).

Lab 3 applied a validated RT-qPCR test procedure on saliva from all 23 positive and a randomly selected set of 86 negatives cases from the 2 cohorts. The demographic results of all 109 saliva samples are mentioned in Table 1.

**Table 1:** Demographic information of patients included in SARS-CoV-2 RT-qPCR testing on saliva

|  |  | # patients | # swab positive | # saliva positive |
| --- | --- | --- | --- | --- |
| age | 5-16 years | 80 | 12 | 13 |
|  | ≥17 years | 29 | 11 | 11 |
| symptoms | yes | 92 | 19 | 20 |
|  | no | 17 | 4 | 4 |
| high-risk contact(s) | yes | 35 | 16 | 17 |
|  | no | 74 | 7 | 7 |

All 23 swab positive samples tested positive using saliva (Table 2, Table 7); in other words, the proportion of saliva-positive samples among swab-positive samples is 100% ([83.1-100.0%] 95% confidence interval), including all swab-positive children’s samples (n=12, [71.8-100.0%] 95% confidence interval, Table 3), all adult samples (n=11, [70.0-100.0%] 95% confidence interval, Table 4), all symptomatic samples (n=19, [80.2-100%] 95% confidence interval, Table 5), and all asymptomatic samples (n=4, [45.4-100%] 95% confidence interval, Table 6).

**Table 2:** 2×2 contingency table of saliva RT-qPCR results versus matched nasopharyngeal or nasal/throat swab results for all cases (n=109)

|  |  | saliva |  |
| --- | --- | --- | --- |
|  |  | + | - |
| swab | + | 23 | 0 |
|  | - | 1 | 85 |

**Table 3:** 2×2 contingency table of saliva RT-qPCR results versus matched nasopharyngeal or nasal/throat swab results for all children (n=80)

|  |  | saliva |  |
| --- | --- | --- | --- |
|  |  | + | - |
| swab | + | 12 | 0 |
|  | - | 1 | 67 |

**Table 4:**
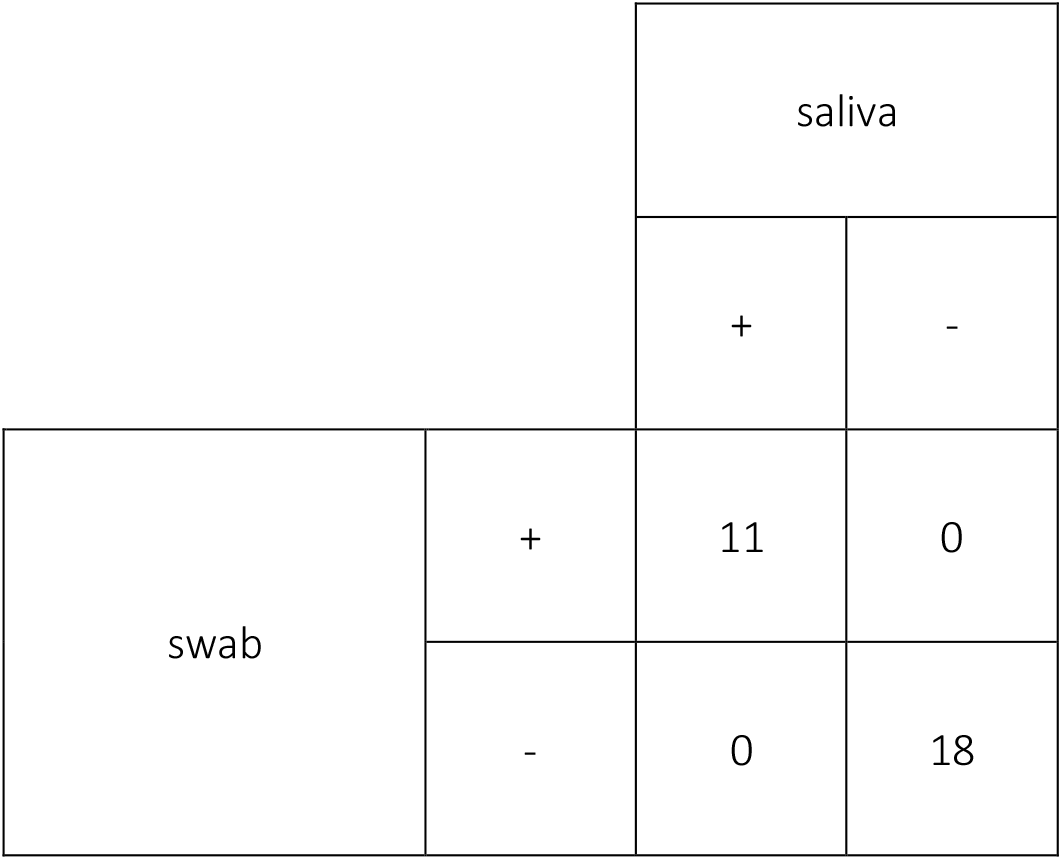
2×2 contingency table of saliva RT-qPCR results versus matched nasopharyngeal or nasal/throat swab results for all adults (n=29)

|  |  | saliva |  |
| --- | --- | --- | --- |
|  |  | + | - |
| swab | + | 11 | 0 |
|  | - | 0 | 18 |

**Table 5:** 2×2 contingency table of saliva RT-qPCR results versus matched nasopharyngeal or nasal/throat swab results for symptomatic cases (n=92)

|  |  | saliva |  |
| --- | --- | --- | --- |
|  |  | + | - |
| swab | + | 19 | 0 |
|  | - | 1 | 72 |

**Table 6:** 2×2 contingency table of saliva RT-qPCR results versus matched nasopharyngeal or nasal/throat swab results for all asymptomatic cases (n=17)

|  |  | saliva |  |
| --- | --- | --- | --- |
|  |  | + | - |
| swab | + | 4 | 0 |
|  | - | 0 | 13 |

**Table 7:** List of patients with positive saliva RT-qPCR result (N/A, not available)

| lab ID | patient ID | saliva Cq | symptoms | high-risk | age | days symptoms | ate/drank | water cleaning mouth | swab Cq (gene) | swab concentration copies/ml | swab type |
| --- | --- | --- | --- | --- | --- | --- | --- | --- | --- | --- | --- |
| 1 | A | 19.0 | yes | yes | 11-16 | N/A | no | N/A | 22.4 (E) | 10<sup>5</sup>–10<sup>7</sup> | nasopharyngeal |
| 1 | B | 19.9 | yes | yes | 5-10 | 1 | no | no | 24.1 (E) | 10<sup>5</sup>–10<sup>7</sup> | nasopharyngeal |
| 1 | C | 22.8 | yes | yes | 5-10 | 2 | no | no | 26.5 (E) | 10<sup>3</sup>–10<sup>5</sup> | nasal/throat |
| 1 | D | 23.8 | yes | yes | 5-10 | N/A | no | N/A | 25.5 (E) | 10<sup>3</sup>–10<sup>5</sup> | nasopharyngeal |
| 1 | E | 27.3 | yes | yes | 5-10 | 1 | no | no | 18.3 (E) | ≥ 10<sup>7</sup> | nasal/throat |
| 1 | F | 31.1 | yes | yes | 5-10 | 7 | yes | no | N/A | N/A | nasopharyngeal |
| 1 | G | 33.3 | yes | yes | 5-10 | N/A | no | no | 33.9 (E) | <10<sup>3</sup> | nasopharyngeal |
| 1 | H | 19.1 | no | yes | 5-10 | N/A | no | no | 27.1 (E) | 10<sup>3</sup>–10<sup>5</sup> | nasopharyngeal |
| 1 | I | 21.6 | no | yes | 11-16 | N/A | yes | no | 23.1 (E) | 10<sup>3</sup>–10<sup>5</sup> | nasopharyngeal |
| 1 | J | 23.6 | no | yes | 5-10 | N/A | yes | no | 25.4 (E) | 10<sup>5</sup>–10<sup>7</sup> | nasal/throat |
| 1 | K | 25.6 | no | yes | 5-10 | N/A | no | no | 23.0 (E) | 10<sup>5</sup>–10<sup>7</sup> | nasopharyngeal |
| 1 | L | 23.3 | yes | no | 5-10 | 1 | no | no | 26.2 (E) | 10<sup>3</sup>–10<sup>5</sup> | nasal/throat |
| 2 | M | 13.9 | yes | yes | 40-49 | 2 | no | yes | 11.2 (N) | N/A | nasopharyngeal |
| 2 | N | 19.0 | yes | no | 17-29 | 2 | no | yes | 17.6 (E) | N/A | nasopharyngeal |
| 2 | O | 19.7 | yes | no | 60-69 | 4 | no | no | 23.0 (E) | N/A | nasopharyngeal |
| 2 | P | 20.2 | yes | yes | 17-29 | 2 | no | no | 9.3 (N) | N/A | nasopharyngeal |
| 2 | Q | 20.8 | yes | no | 30-39 | 1 | no | no | 13.6 (N) | N/A | nasopharyngeal |
| 2 | R | 24.1 | yes | yes | 30-39 | 1 | yes | no | 12.2 (N) | N/A | nasopharyngeal |
| 2 | S | 24.4 | yes | no | 17-29 | 1 | no | no | 15.9 (N) | N/A | nasopharyngeal |
| 2 | T | 24.8 | yes | yes | 30-39 | 1 | no | yes | 31.4 (E) | N/A | nasopharyngeal |
| 2 | U | 25.5 | yes | no | 17-29 | 3 | no | yes | 29.5 (E) | N/A | nasopharyngeal |
| 2 | V | 28.4 | yes | yes | 11-16 | 2 | no | no | 18.7 (N) | N/A | nasopharyngeal |
| 2 | W | 32.2 | yes | yes | 30-39 | 2 | no | no | 24.9 (E) | N/A | nasopharyngeal |
| 2 | X | 33.8 | yes | no | 50-59 | N/A | no | yes | 31.9 (E) | N/A | nasopharyngeal |

Saliva from the 86 swab-negative patients was confirmed to be negative for all but one sample. One saliva sample from a child was tested low positive (Cq=31.1, Table 7), 7 days after developing COVID-19 symptoms. Hence, the relative sensitivity of RT-qPCR saliva testing was 104.3% across all patients.

While direct quantitative comparison of Cq values across laboratories is not recommended, the Cq values between swab and saliva are largely comparable, with a median difference of 1.9 cycles in favor of the swab result (Figure 2).

**Figure 2:**
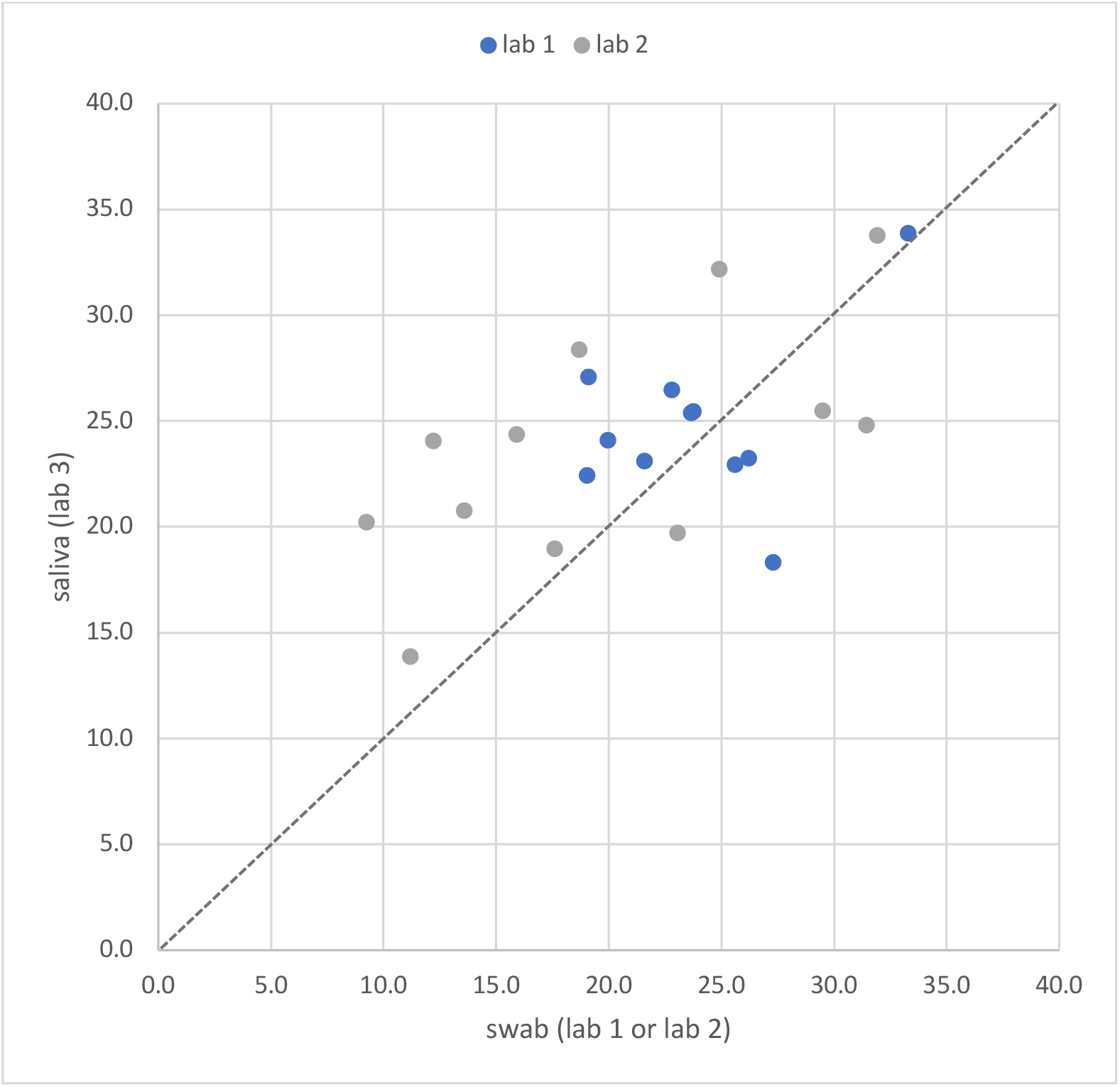
Cq of matched nasopharyngeal or nasal/throat swab results (x-axis, lab 1 or lab 2) vs. saliva result (y-axis, lab 3)

To assess the impact of eating/drinking or rinsing the mouth prior to saliva collection on the SARS-CoV-2 RT-qPCR detection sensitivity, we compared the difference in Cq value of the spike-in RNA control (as a measure for inhibition) between these 2 groups, or the difference between the saliva Cq and the swab Cq value (delta-Cq) for SARS-CoV-2 between these groups. Both analyses provide no evidence that eating or drinking 30 minutes prior to saliva collection, or rinsing the mouth with water 10 minutes prior to saliva collection negatively affect SARS-CoV-2 detection sensitivity (p-values > 0.05). With the smallest group size being 18 and an observed standard deviation of spike-in RNA Cq of 0.375, we had >95% power to detect a 0.5 cycle difference.

## Discussion

PCR-based testing for SARS-CoV-2 has been instrumental in the global effort to control the COVID-19 pandemic. While nasopharyngeal swabs are widely recommended to maximize detection sensitivity, this sampling procedure comes with significant discomfort, especially for children, and requires trained staff for collection. Furthermore, maximizing diagnostic sensitivity may not be the best strategy to prevent spreading; instead, frequency of testing should be prioritized over sensitivity in controlling the spread of this virus^5^. Saliva may provide an excellent alternative for a swab as it allows non-invasive and repeated self-collection and has been demonstrated to result in equivalent sensitivity^3,4^. Nonetheless, noticeable performance differences among individual studies are published, likely resulting from varying collection devices, with or without stabilizing medium (likely important because of large amounts of RNases in saliva), sample storage conditions, time delays between collection and testing, phase of the pandemic^6^ during which sampled are collected, donor inclusion criteria (hospitalized vs. asymptomatic persons), and unstandardized laboratory saliva testing. Also, different ways of saliva collection are reported, including spitting (either or not stimulated), gargling, or posterior oropharyngeal spitting (throat clearing), and varying recommendations to refrain from eating or drinking, and rinsing the mouth prior to collection.

In our study, we used a novel saliva collection device for supervised self-collection of a small volume of 1.3 ml unstimulated saliva in a stabilizing buffer that inactivates infectious agents and stabilizes RNA. A small volume of saliva is an important benefit, especially for children and elderly people, who have great difficulties in producing large saliva volumes. Most published studies require at least 2 and up to 5 ml of saliva. In our study, we included children from the age of 5 years onwards as they can easily produce a 1.3 ml spitting sample, an age cut-off also recommended by Delaney et al.^7^.

While our patient cohort size is modest, our results are perfectly in line with recent meta-analyses on the use of saliva as an alternative to nasopharyngeal swabs^3,4^. We observed a 100% concordance, across all demographic groups, irrespective of age, presence of symptoms, or high-risk status. Our study has not observed any false negatives, and -despite the difficulties to compare Cq values across laboratories-general good concordance in Cq values between saliva and swab. In line with previous reports (reviewed in ^3^), we have detected one case that is saliva positive and swab negative. Of note, this child was sampled 7 days after symptoms started, the longest period in our cohort. It remains to be determined whether the higher relative sensitivity observed for saliva is due to variation in nasopharyngeal sampling^10^ or due to differential viral load dynamics over time in function of body part^11^.

While neat saliva may pose handling challenges because of its complex matrix with non-Newtonian behavior and high viscosity, we did not observe any pipetting problems in our study. One possible explanation may be the reported mucolytic effect of some components of the InActiv Blue transport medium, such as guanidine thiocyanate and sarkosyl^8,9^. We did also not observe any signs of RT-qPCR inhibition or loss of sensitivity when comparing patients with respect to their eating/drinking or mouth rinsing behavior prior to saliva collection. Together with the 100% concordance rate, our results therefore suggest that the reported recommendations to refrain from drinking or eating 30 minutes prior to saliva collection or rising the mouth with water 10 min prior to collection may not be universally valid.

While not specifically tested in this study, saliva also holds promise to detect other respiratory viruses, like RSV and influenza^12^. This may be of great value for differential diagnosis of SARS-CoV-2, RSV and influenza using the same saliva sample.

## Materials and Methods

The study was approved by the Ghent University Hospital ethics committee (B6702021000459) for parallel collection of saliva from children aged 5-16 and adults during a visit at the general practitioner (GP) during which a swab is collected for diagnostic purposes. At the same time, for each patient, a short survey is completed to enquire about symptoms, high-risk contacts, and eating/drinking behavior or mouth rinsing with water prior to saliva collection.

At the GP, saliva was collected using the commercially available CE marked Saliva Collection Kit (InActiv Blue, IB_COL) according to the kit’s instructions (∼1.3 ml saliva + 2 ml InActiv Blue^®^) and a nasopharyngeal (lab 1, lab 2) or combined nasal/throat swab (lab 1) was collected in 2 ml VST medium (#456162, Greiner Bio-One; lab 1) or in 2 ml of InActiv Blue^®^ (#456604, InActiv Blue; lab2). InActiv Blue^®^ is a virus inactivating and RNA stabilizing buffer that protects RNA for up to 30 days at room temperature. Sample transport from the GP to medical lab 1 or 2 was performed at room temperature, followed by immediate processing of the swab sample according to the routine diagnostic procedure. The saliva samples were stored at 2-8 °C (lab 1) or frozen (lab 2) and shipped to lab 3 for further testing.

Upon arrival at lab 3, the samples were processed according to an ISO 17025 accredited procedure. The saliva samples were first thawed, and the tubes were put in an oven (Binder FP115) at 83 °C for a period of 10 to 20 minutes, depending on number of tubes, such that the cap reaches 70 °C for at least 5 minutes (heat camera verified). The heat procedure does not only guarantee complete inactivation of the thread of the screw cap (not exposed to the inactivating buffer) but may also help render the sample less viscous. Hundred μl of saliva was aspirated using a Tecan Freedom EVO 200 liquid handler, followed by MagSI-NA Pathogens RNA extraction (magtivio #MDKT00210960) on a PurePrep 96 instrument (magtivio) and eluted in 75 μl. Six μl of RNA eluate was used as input for a 20 μl duplex RT-qPCR reaction in a CFX384 qPCR instrument (Bio-Rad) using 10 μl One Step PrimeScript III (Takara Bio #RR600B) according to the manufacturer’s instructions, and 250 nM final concentration of primers and 400 nM of hydrolysis probe. Primers and probes were synthesized by Integrated DNA Technologies using cleanroom GMP production. For detection of the SARS-CoV-2 virus, the Charité E-gene^13^ and CDC N2 gene primers/probe^14^ were used (both in FAM channel); for the internal spike-in control, a proprietary hydrolysis probe assay (HEX channel) was used. Cq values were generated using the FastFinder software v3.300.5 (UgenTec).

In medical lab 1 (Labo Nuytinck), nasopharyngeal or nasal/throat samples were analyzed using a validated routine RT-qPCR test (Allplex SARS-CoV-2 Assay; Seegene, Accuramed), consisting of RNA extraction with STARMag 96X4 viral DNA/RNA 200C kit (Seegene, Accuramed) on a Hamilton Starlet followed by RT-qPCR on an CFX96 qPCR instrument (Bio-Rad). Seegene viewer v3 was used for amplification curve interpretation. For positive samples, the Cq values of E, RdRP/S and N genes were reported. Semi-quantitative swab viral concentration was calculated using reference material provided by the Belgian Reference Center.

In medical lab 2 (Medisch Labo Bruyland), nasopharyngeal samples were analyzed using a validated routine RT-qPCR test. Most of the samples were analyzed (n = 21) using the ThermoFisher platform (reporting N, S and ORF genes). Prior to the analysis, the nasopharyngeal samples were heat-inactivated and were transferred into (600 µl) transparent tubes. RNA was extracted from 200 µl sample using MagMAX Viral/Pathogen II Nucleic Acid Isolation kit (ThermoFisher), Tecan Freedom EVO 100 liquid handler (Tecan) and KingFisher Flex (ThermoFisher). The barcoded PCR plate was prepared using the RNA extracts, TaqPath COVID-19 CE-IVD RT-PCR kit (ThermoFisher) and Tecan Freedom EVO 100 liquid handler. PCR was carried out on a QuantStudio5 (ThermoFisher). Data were analyzed using the FastFinder software v4.5.2 (UgenTec). For the SARS-CoV-2 positive samples, the Cq values of N, S and ORF genes were reported. A minority of the samples (n = 15) was analyzed using the Roche Platform (reporting E gene). Prior to the analysis, the samples were transferred to flow tubes (appropriate for the Roche platform) and an equal volume of Cobas PCR Medium was added to inactivate the samples. RNA was extracted using the Flow primary sample handling pipetting robot (Roche/Hamilton), MagNA Pure 96 DNA and Viral NA Small Volume kit (Roche) and MagNA Pure 96 (Roche). The barcoded PCR plate was prepared using the RNA extracts, RNA process Control kit (Roche), LightMix Modular Sarbecovirus SARS-CoV-2 (Roche/TIB Molbiol) and Flow PCR Setup pipetting robot (Roche/Hamilton). PCR was performed using the LightCycler 480 II. Data were analyzed using the FLOW software (Roche) and for the positive samples, the E gene Cq values were reported.

Cq values used in figures or mentioned in tables are E gene for lab 1, E or N gene for lab 2, and combined E/N gene for lab 3.

This relative sensitivity was calculated by dividing the sensitivity of testing on saliva by the sensitivity on swabs. The relative sensitivity can take values from zero to infinity. A value above one indicates that testing on saliva is more sensitive than on the swab. Confidence intervals on proportions of counts were calculated using GraphPad’s QuickCalcs according to the modified Wald method. An unpaired t-test (using MS Excel version 16.52) was used to compare Cq values of the spike-in RNA between saliva samples from patients who either did or not eat/drink 30 minutes prior to saliva collection, or who either did or did not rinse their mouth with water 10 minutes prior to saliva collection. The same test was used to compare delta-Cq values (saliva Cq – swab Cq for SARS-CoV-2 positive cases) between the aforementioned groups. Power analysis for a t-test was calculated using Piface version 1.76.

## Data Availability

All relevant data is provided in the Tables.

## Acknowledgements

We are grateful to all patients, parents, and general practitioners for sample collection. We also want to thank all lab technicians for their dedicated work during this study, and Michiel Vandewalle for making Figure 1. We would like to thank Katrien Vandewiele for the recruitment of the medical doctors and Liesbet Demaegd and the MLTs from the COVID-team for the analysis of the samples.

